# Lower Urinary Tract Symptoms in a prospective cohort of COVID-19 survivors

**DOI:** 10.1101/2023.08.01.23293500

**Authors:** Cristiano M. Gomes, Marcelo Hisano, Julia D. Souza, João Victor T. Henriques, Jose de Bessa, Julyana Moromizato, Thulio Bosi, Rachel Mazoni, João Gismondi, Bruno Camargo, Zein M. Sammour, Homero Bruschini, Linamara R. Battistella, William C. Nahas, the HCFMUSP COVID-19 Study Group

## Abstract

**Purpose:** To analyze the prevalence of lower urinary tract symptoms (LUTS) in patients who survived moderate and severe forms of COVID-19 and the risk factors for LUTS six months after hospital discharge.

**Materials and Methods:** In this prospective cohort study, patients were evaluated six months after being hospitalized due to COVID-19. LUTS were assessed using the International Prostate Symptom Score. General health was assessed through the Hospital Anxiety and Depression Scale and the EQ5D-L5 scale, which evaluates mobility, ability to perform daily activities, pain and discomfort and completed a self-perception health evaluation.

**Results:** Of 255 participants, 54.1% were men and the median age was 57.3 [44.3 – 66.6] years. Pre-existing comorbidities included diabetes (35.7%), hypertension (54.5%), obesity (30.2%) and physical inactivity (65.5%). 124 (48.6%) had a hospital stay >15 days, 181 (71.0%) were admitted to an ICU and 124 (48.6%) needed mechanical ventilation. Median IPSS score was 6 [3-11] and did not differ between men and women. Moderate to severe LUTS affected 108 (42.4%) patients (40.6% men and 44.4% women; p=0.610). Nocturia (58.4%) and frequency (45.9%) were the most prevalent symptoms and urgency was the only symptom that affected men (29.0%) and women (44.4%) differently (p=0.013). LUTS significantly impacted the quality of life of 60 (23.5%) patients with women more severely affected (*p*=0.004). Preexisting diabetes, hypertension and self-perception of worse general health were associated with LUTS.

**Conclusions:** LUTS are highly prevalent and bothersome six months after hospitalization due to COVID-19. Assessment of LUTS may help ensure appropriate diagnosis and treatment in these patients.

## Introduction

The COVID-19 pandemic has had a devastating impact on public health, resulting in millions of hospitalizations and deaths globally.^1^ Among survivors, post-acute sequelae of SARS-CoV-2 are frequent and a major cause of concern, as it overloads health systems worldwide.^2,3^ This syndrome may also been referred to as “long COVID”.^4^ Studies conducted in different countries have shown persistent multiple organ manifestations including pulmonary dysfunction, muscle weakness and fatigue, neurological and psychological disorders, cardiovascular disorders, digestive disorders and other unspecific symptoms such as hair loss, in up to 76% of COVID-19 survivors four to six months after hospital discharge.^5-7^

COVID-19 can be associated with *de novo* lower urinary tract symptoms (LUTS), such as urgency, urinary frequency and nocturia, or it can deteriorate pre-existing LUTS during the acute phase.^8-11^ However, the long-term effects of COVID-19 on LUTS have been poorly evaluated and the prevalence of LUTS in patients who recovered from COVID-19 remains unknown. In a study from Greece, a high prevalence of increased urinary frequency and urgency consistent with the diagnosis of overactive bladder has been reported in patients who were suffering with long COVID.^14^ The study did not evaluate the prevalence of LUTS in the population, since they only included 66 patients who reported having urinary urgency. Moreover, they did not state how long after acute COVID the patients were evaluated. In another study consisting of a small case series, persistent LUTS and urodynamic abnormalities have been shown months after acute SARS-CoV-2 infection in teenagers.^15^ Authors proposed that clinicians should be aware of a recent COVID-19 infection in patients with sudden onset lower urinary tract dysfunction.

The aim of the present study was to analyze the prevalence of LUTS in men and women who survived moderate and severe forms of COVID-19. We also evaluated the potential impact of preexisting comorbidities and clinical parameters of COVID-19 severity on the prevalence of LUTS as well as the association of LUTS with other symptoms and conditions related to Long COVID-19. To this end, we utilized a large, hospital-based dataset to identify a cohort in the acute phase of the disease, and then completed a longitudinal follow-up of this cohort.

## Materials and Methods

This is a cohort study conducted at a university based, tertiary medical facility that provided care for moderate and severe cases of the COVID-19 during the acute phase of the first wave of the pandemic, i.e., prior to the onset of vaccination protocols. This cohort was constituted to facilitate multidisciplinary studies addressing long-term medical and functional outcomes among adults who survived COVID-19. Details about the methodological protocol can be found elsewhere.^16^

Participants: All eligible adult subjects (≥18 years) who had been hospitalized in our hospital for treatment of moderate or severe COVID-19 for at least 24 hours and had laboratory-confirmed SARS-CoV-2 infection were invited to participate.

Assessment protocol: Participants underwent a one-to-two days series of multidisciplinary evaluations performed six months after hospital discharge including thorough, multi-domain questionnaires applied by medical research staff, self-report scales, objective evaluations of cardiopulmonary functioning and physical functionality. The study was approved by the local ethics committee (approval number 4.270.242). Informed consent was provided by all participants.

### Preexisting comorbidities

The evaluated preexisting comorbidities were diabetes mellitus, systemic arterial hypertension (SAH), obesity (body mass index >30) and physical inactivity (i.e., < 150 min/week at moderate-to-vigorous physical activity)

### Parameters of COVID-19 severity

Data regarding the acute stage of SARS-CoV-2 infection was retrieved from hospital charts and databases, providing information on length of hospital stay, admission to an intensive care unit (ICU), need for invasive mechanical ventilation and need for hemodialysis.

### Prevalence of LUTS six months after hospital discharge

LUTS were assessed using the International Prostate Symptom Score (IPSS) questionnaire.^17^ Participants were asked to rate how often they experienced individual LUTS during the past month. It was used to analyze the presence of individual lower urinary tract symptoms of frequency, incomplete emptying, intermittency, urgency, slow stream, straining to void and nocturia (≥ 2 episodes/night). We considered each symptom to be present when it occurred less than half the time or more.^18, 19^ LUTS were considered mild when IPSS score < 7, moderate if IPSS between 8 and 19 and severe when IPSS ≥ 20. The impact of LUTS on quality of life (QoL) was evaluated on a scale of 0 (“delighted”) to 6 (“terrible”). A significantly impaired QoL was defined for patients who were mostly dissatisfied or worse.

A urinalysis was obtained at the six months evaluation and was considered normal if there were ≤ 10 leukocytes per field and ≤ 3 red blood cells per field.

### General health six months after hospital discharge

To evaluate the overall health status after 6 months of hospital discharge, patients completed a series of evaluations. Mental health was assessed through the Hospital Anxiety and Depression Scale (HADS).^20^ Patients’ self-rated health was assessed through the EQ5D-L5 scale, which evaluates mobility, ability to perform personal care and daily activities, pain, discomfort and anxiety/depression.^21^ Additionally, we asked patients to rate their self-perception of overall health on a Likert scale varying from very good to good, reasonable, bad or very bad. An impaired health status was considered when patients rated themselves as bad or very bad.

### Statistical analysis

Quantitative variables were expressed as medians and interquartile ranges, while qualitative variables were expressed as absolute values, percentages, or proportions. Student’
ss t or ANOVA was used to compare continuous variables. Categorical variables were compared using the Chi-squared or Fisher’s exact test. Associations were described as Odds Ratios with respective confidence intervals. Correlations were analyzed by Pearson’s correlation coefficient.

All tests were 2-sided, and a p-value < 0.05 was considered statistically significant. Analysis was performed using commercially available statistical software (GraphPad Prism, version 9.03 for Windows, San Diego California USA).

## Results

Of 1,987 eligible patients invited to participate in the study, 870 agreed to participate in the multidisciplinary assessment. Because the evaluation of LUTS was included when the study was ongoing for some time, the cohort of patients included for LUTS assessment included only 255. Men comprised 54.1% of study participants, and the median age of the participants was 57.3 [44.3 – 66.6] years.

### Pre-existing comorbidities

Pre-existing comorbidities included diabetes mellitus (35.7%), hypertension (54.5%), obesity (30.2%) and physical inactivity (65.5%). Men and women were comparable in terms of diabetes, hypertension and physical activity habits, but obesity was more common among women (26.8% vs 34.2%; p=0.026 - Table 1).

**TABLE 1.**
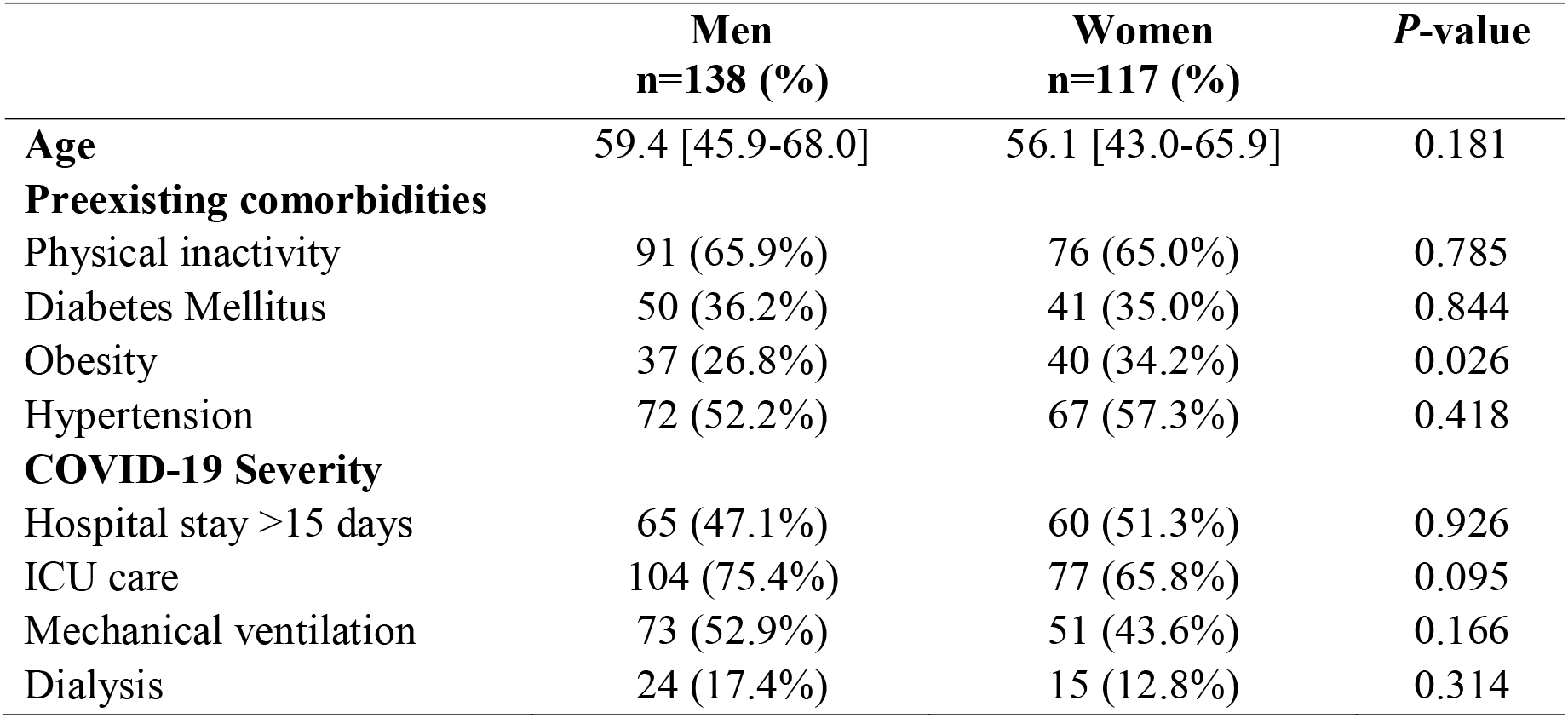
Demographic characteristics, preexisting comorbidities and COVID-19 severity in the study population (N=255)

### COVID-19 severity parameters

Of the 255 participants, 124 (48.6%) had a hospital stay >15 days, 181 (71.0%) were admitted to an ICU, 124 (48.6%) needed mechanical ventilation and 39 (15.3%) needed hemodialysis. Men and women did not differ in terms of COVID-19 severity parameters (Table 1).

### Prevalence of LUTS 6 months after hospital discharge

The median IPSS score was 6 [3-11] and did not differ between men and women (7.0 [3.0-11.0] and 5.0 [2.0-11.0], respectively; *p*=0.371). Moderate to severe LUTS affected 108 (42.4%) patients (40.6% men and 44.4% women; p=0.610). Age was not correlated with total IPSS scores (r=0.099, p=0.109) and was similar between patients with mild vs moderate to severe LUTS (56.3 [42.8-66.5] vs 58.5 [47.2-66.75] respectively; p=0.341).

Nocturia (58.4%) and increased urinary frequency (45.9%) were the most prevalent symptoms. Urgency was the only symptom that affected men (29.0%) and women (44.4%) differently (p=0.013). Storage symptoms were more common in women (5.0 [3.0-7.0] vs 3.0 [1.0-7.0]; p=0.024), while voiding symptoms affected both genders similarly (2.0 [0-4.0] vs 1 [0-4.8]; p=0.641) (Table 2).

**TABLE 2.**
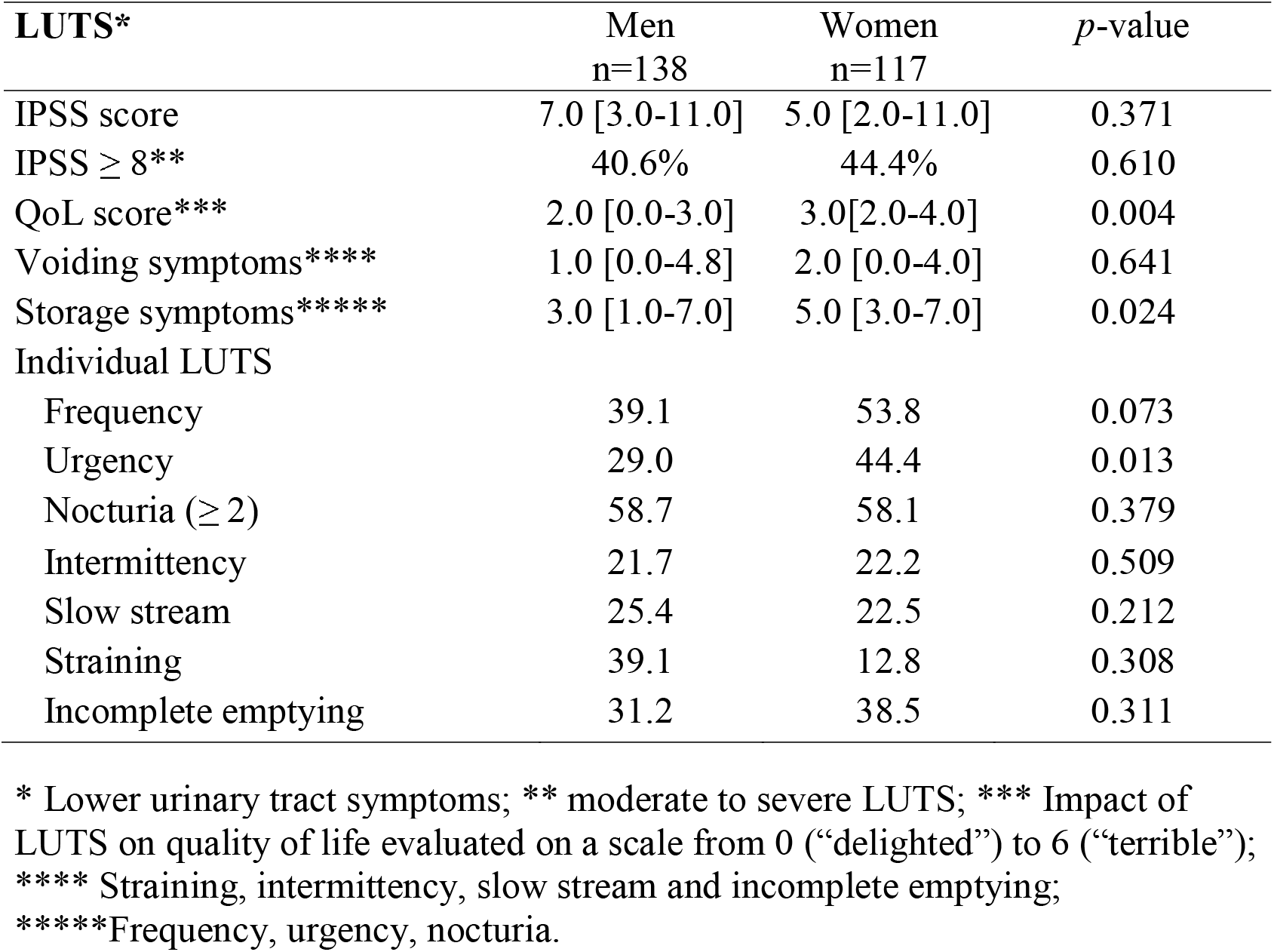
Prevalence of LUTS six months after COVID-19.

LUTS significantly impacted the QoL of 60 (23.5%) patients. Women were more severely affected with a median QoL score of 3 [2-4] vs 2 [0-3] (*p*=0.004) (Table 2).

Urinalysis obtained from 229 (89.8%) participants at the six-month evaluation was normal in 193 (84.3%) patients while 27 (11.8%) had leukocyturia, 13 (5.7%) had hematuria and 4 (1.7%) had both. The severity of LUTS did not differ between patients with normal and abnormal urinalysis (p=0.526).

### General health six months after hospital discharge

According to the HADS score, the mean anxiety subscore was 5.64 ±4.83 with worse scores among women (7.70±5.05 vs 3.85±3.82; *p*<0.001). Sixteen (11.6%) men and 44 (37.6%) women had a HADS anxiety subscore ≥ 8, which is consistent with possible anxiety disorder. The mean depression subscore was 4.56 ±4.41 and women had higher scores (5.71±4.57 vs 3.55±4.01; p<0.001). Seventeen (12.3%) men and 33 (28.2%) women had a HADS depression subscore ≥ 8, which is consistent with depression.

Based on the EQ-5D questionnaire, 27 (19.6%) men and 36 (30.8%) women reported moderate to severe difficulty to walk, 13 men (9.4%) and 14 women (12.0%) reported moderate to severe difficulty to wash and dress themselves and 22 men (15.9%) and 35 women (29.9%) reported moderate to severe problems to do their usual activities. Forty-four men (31.9%) and 64 women (54.7%) had moderate to strong pain or discomfort. Twenty-seven men (19.6%) and 61 women (52.1%) considered themselves to be moderately to extremely anxious or depressed.

On the Likert scale, 29 (11.4%) patients considered their health as bad or very bad, including 17 (14.5%) women and 11 (8.0%) men (p=0.073).

### Association of LUTS, comorbidities, COVID severity, general health and anxiety/depression

Moderate to severe LUTS were more prevalent among individuals with preexisting diabetes (*p*=0.025) and SAH (*p*<0.001). No associations were found between LUTS and parameters of COVID-19 severity. Among the parameters of general health 6 months after hospital discharge only impaired overall health was associated with LUTS (*p*=0.026) (table 3).

**TABLE 3.**
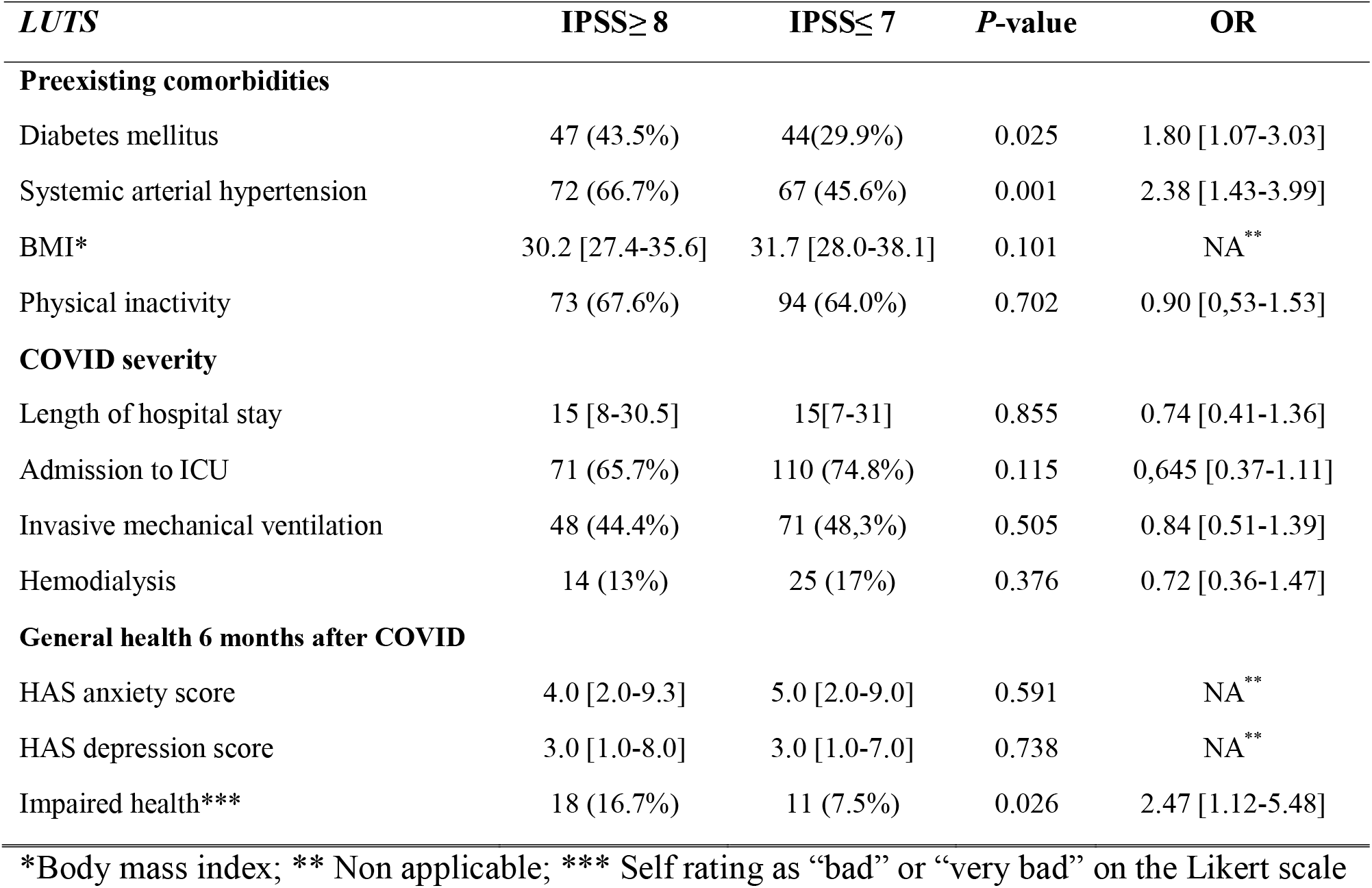
Association of LUTS severity with preexisting comorbidities, COVID severity and general health parameters six months after hospital discharge.

## Discussion

To our knowledge, this is the first prospective cohort study of hospitalized COVID-19 survivors to date that has focused on the assessment of LUTS. We showed a high prevalence of LUTS in this population, with 42.4% reporting moderate to severe LUTS. Storage symptoms were more common in women, while voiding symptoms affected both genders similarly. LUTS significantly impacted the QoL of 23.5% of the patients and women were more severely affected. Age was not correlated with symptom scores and was similar between patients with mild vs moderate to severe LUTS. Preexisting diabetes, hypertension and having an impaired overall health six months after COVID were associated with higher rates of moderate to severe LUTS. No associations were found between LUTS and parameters of COVID-19 severity.

Long COVID may include an array of symptoms and diseases, but LUTS have not been included as part of the symptom profile of long COVID. Our results showing that 42.4% of the patients had moderate to severe LUTS seem to highlight the relevance of LUTS in this population. We used the IPSS to evaluate LUTS. It is a widely used tool to assess LUTS, which has commonly been used in epidemiological studies with mixed-gender populations.^19, 22, 23^ The prevalence of moderate-to-severe LUTS in Brazil has been reported in 20.8% of men and 23.9% of women, which is similar to other countries and roughly half the rates observed in our population of long COVID subjects with similar age distribution.^18, 19^ We showed a predominance of storage symptoms in patients with long COVID, which is consistent with studies reporting on LUTS in the acute phase of COVID-19^8, 11, 23^ and also with the study from Zachariou et al, who found a high prevalence of overactive bladder among patients who had been hospitalized due to COVID-19.^14^

Interestingly, we found minor differences between men and women and no impact of age in the prevalence of LUTS. This negative finding might be explained by our patient sample, which was relatively small. Second, critical illness due to multiorgan disease including psychiatric (depression and anxiety), fatigue, neuromuscular and pulmonary symptoms might attenuate the effect of aging.^24^

It is known that acute COVID-19 infection can cause LUTS, although the mechanism is not understood. A systematic review found that SARS-CoV-2 can possibly damage the prostate and worsen LUTS due to BPH.^12^ The proposed mechanism is that SARS-CoV-2 binds to angiotensin converting enzyme 2 (ACE2) receptors causing downregulation of ACE2, which might trigger inflammation and progression of BPH. Another possible mechanism of LUTS is due to neurological disorders, including peripheral nervous system involvement.^13^ In long COVID, LUTS are probably multifactorial and associated with the increased comorbidity profile of the patients. There is no evidence that SARS-CoV-2 may have a direct effect in the lower urinary tract leading to long term LUTS. However, several risk factors or complications of long COVID could contribute to LUTS including age, obesity, diabetes and metabolic disorders, cardiovascular and lung diseases, neurological conditions, anxiety, depression and sleep disorders.^25-28^ In our study, diabetes and hypertension were risk factors for LUTS as well as having an impaired overall health six months after hospital discharge. Parameters of COVID severity such as need for ICU or mechanical ventilation were not associated with the prevalence of LUTS.

The impact of LUTS in our population was significant, with 23.5% reporting that they were mostly dissatisfied or worse with LUTS. It is well known that LUTS may have a significant impact on patients’ QoL causing physical discomfort, emotional distress, and disruptions in daily activities.^25, 29^ Combined with the lingering effects of long COVID, LUTS may further exacerbate the challenges faced by individuals in their recovery process.^28^ Moreover, LUTS can impact mental well-being and exacerbate anxiety or depression symptoms.^25^ Finally, LUTS, such as urgency and frequency, can cause functional limitations by affecting mobility and social activities.^24^ These findings highlight the importance of healthcare providers being aware of the potential association between long COVID and LUTS.

The COVID-19 landscape is continually changing across the world. In the past three years, we have witnessed the rise and spread of multiple variants, while vaccination programs advanced in multiple countries. All of the subjects included in this study had COVID between March and August 2020, when vaccines weren’t yet available and the B.1.1.28 and B.1.1.33 lineages were the most prevalent in Brazil. Future studies should evaluate differences among the variants of the SARS-CoV-2 as well as the effect of vaccination.

Our study has several limitations. First, we cannot extrapolate our findings to other countries or communities, since conditions may vary in terms of SARS-CoV-2 variant, vaccination status and overall epidemiology. Also, our study sample is relatively small, and we had no control group for comparisons. Another shortcoming is the fact that we do not have data regarding patients’ previous LUTS.

We believe our findings have significant implications for clinical practice, informing urologists and other health care professionals involved in the management of patients with LUTS that a history of moderate or severe COVID-19 requiring hospitalization may increase the risk of LUTS. Second, our findings should be important to healthcare professionals treating patients with long COVID since LUTS may have a significant impact on patients’ QoL and is certainly a neglected aspect of their health. Thus, our results should encourage the development of multidisciplinary approaches to manage long COVID recovery.

## Conclusions

LUTS are highly prevalent and often bothersome six months after hospitalization due to COVID-19. Future studies should clarify the role of different virus variants and vaccination on the prevalence of long COVID and LUTS. Comprehensive assessment of LUTS and their effects may help ensure appropriate diagnosis and treatment in these patients.

## Data Availability

All data produced in the present study are available upon reasonable request to the authors

## Acknowledgments

The authors would like to thank the participants of the study for their time.

